# Pre-Omicron vaccine breakthrough infection induces superior cross-neutralization against SARS-CoV-2 Omicron BA.1 than primo infection

**DOI:** 10.1101/2022.06.21.22276659

**Authors:** Eveline Santos da Silva, Michel Kohnen, Georges Gilson, Therese Staub, Victor Arendt, Christiane Hilger, Jean-Yves Servais, Emilie Charpentier, Olivia Domingues, Chantal J. Snoeck, Markus Ollert, Carole Seguin-Devaux, Danielle Perez-Bercoff

**Affiliations:** HIV Clinical and Translational Research Unit, Department of Infection and Immunity, Luxembourg Institute of Health, 29 rue Henri Koch, L-4354 Esch-sur-Alzette, Luxembourg; Centre Hospitalier de Luxembourg, 4 rue Ernest Barblé, L-1210 Luxembourg, Luxembourg; Molecular and Translational Allergology, Department of Infection and Immunity, Luxembourg Institute of Health, 29 rue Henri Koch, L-4354 Esch-sur-Alzette, Luxembourg; Clinical and Applied Virology, Department of Infection and Immunity, Luxembourg Institute of Health, 29 rue Henri Koch, L-4354 Esch-sur-Alzette, Luxembourg; Allergy and Clinical Immunology, Department of Infection and Immunity, Luxembourg Institute of Health, 29 rue Henri Koch, L-4354 Esch-sur-Alzette, Luxembourg; Odense Research Center for Anaphylaxis, Department of Dermatology and Allergy Center, Odense University Hospital, University of Southern Denmark, 5000 Odense, Denmark

## Abstract

SARS-CoV-2 variants raise concern because of their high transmissibility and their ability to evade neutralizing antibodies elicited by prior infection or by vaccination. Here we compared the ability of sera from 70 unvaccinated COVID-19 patients infected before the emergence of variants of concern (VOCs) and from 16 vaccine breakthrough infection (BTI) cases infected with Gamma or Delta to neutralize the ancestral B.1 strain, and the Gamma, Delta and Omicron BA.1 variants using live virus. We further determined antibody levels against the Spike protein, the Receptor Binding Domain (RBD) and the N-terminal domain (NTD) of Spike. Convalescent sera featured considerable variability in neutralization of B.1 and in cross-neutralization of different strains, and neutralizing capacity moderately correlated with antibody levels against Spike and the RBD. All but one convalescent serum failed to neutralize Omicron BA.1. Overall, convalescent sera from patients with moderate disease had higher antibody levels and displayed higher neutralizing ability against all strains than patients with mild or severe forms of disease. Sera from BTI cases fell into one of two categories: half the sera had high neutralizing activity against the ancestral B.1 strain as well as against the infecting strain, while the other half had no or very low neutralizing activity against all strains. Although antibody levels against Spike and the RBD were lower in BTI cases than in unvaccinated convalescent sera, most neutralizing sera also retained partial neutralizing activity against Omicron BA.1, indicative of cross-neutralization between B.1, Delta and Omicron and suggestive of higher affinity, as confirmed by the IC50:Ab level ratios. Neutralizing activity of BTI sera was strongly correlated with antibodies against Spike and the RBD. Together, these findings highlight qualitative differences in antibody responses elicited by infection in vaccinated and unvaccinated individuals. They further suggest that breakthrough infection with a pre-Omicron variant boosts immunity and induces cross neutralizing antibodies against different strains, including Omicron BA.1.

## Introduction

Two years into the COVID-19 pandemic which originated in Wuhan, China, in December 2019, SARS-CoV-2 has officially infected over 534 million individuals and claimed more than 6.31 million lives as of June 16^th^ 2022. SARS-CoV-2 variants continuously mold the pandemic landscape. Variants that spread faster or elude immunity conferred by prior infection or by vaccines readily outcompete established strains. The first evolution of the Wuhan strain (B.1) emerged between mid-February and mid-March 2020 [1-3]. It harbors the D614G mutation in Spike (S-D614G) which favors viral infectivity and transmission by positioning Spike in an ‘up’ position prone to bind ACE-2 [4]. Numerous other lineages have stemmed since from B.1, including the main variants of concern (VOCs) Alpha (B.1.1.7), Beta (B.1.351), Gamma (P.1) and Delta (B.1.617.2). Aside from increased transmission, infection with the Alpha and Delta variants is associated with higher disease severity and fatality rate [5-14], including in children and adolescents [15, 16]. The recent Omicron (B.1.1.529) variants BA.1 and BA.2, which were first identified in Sub-Saharian Africa in November 2021, have supplanted Delta within a few weeks causing two overlapping peaks in most countries [17]. Their infectivity and transmissibility are far higher than Delta and yet more transmissible sub-lineages are now blossoming. These variants are in turn being outgrown by BA.2 sub-variants (notably B.1.12.1) and BA.4 and BA.5 [18, 19]. Omicron variants cause less severe forms of COVID-19 (∼60% lower risk of hospitalization or death compared to Delta) [20-23], reflecting the combined effects of intrinsic lower viral pathogenicity and prior immunity [24-28]. The large number of deletions and mutations (> 60 across the genome, including 37 in the viral protein Spike and R203K in the Nucleocapsid N), improve its affinity for the viral receptor ACE2 on target cells [21, 29-31] and thereby its transmissibility, accelerate viral assembly and enable immune evasion [32-36].

Encapsulated mRNA-based vaccines BNT162b2 (Pfizer-BioNTech) and mRNA-1273 (Moderna) as well as Adenovector-based vaccines ChAdOx1 (Astra Zeneca, AZ) and Ad26.Cov2.S (Janssen) confer effective protection against severe forms of COVID-19 [10, 37-41]. Protection against infection and transmission is less striking, particularly for the two latest VOCs Delta and Omicon [10, 39-50], as testified by increasing numbers of reinfection and breakthrough infections [27, 33, 38, 51-58]. The waning of immune responses and the emergence of variants with mutations in Spike [33-35, 37-40, 43, 48, 49, 51, 52, 59-71] are the main reasons of vaccine escape and reinfections.

Convalescent patient sera and sera from individuals vaccinated with mRNA vaccines BNT162b or mRNA-1273 have lower *in vitro* neutralizing activity against VOCs than against B.1. For Alpha and Delta, the drop in neutralizing activity is modest (2-3-fold) [48, 72-77], while it reaches ∼10-fold to full escape for Beta and Gamma [48, 72-79]. In agreement with *in vitro* tests, protection conferred by the BNT162b2 and the AZ ChAdOx1 vaccines against the Alpha variant is only moderately decreased [37, 60, 62, 64]. Protection against the Delta variant dropped slightly, from > 95% to 58-83% for BNT162b2 and from 70 to 60% for AZ ChAdOx1 [38, 51-53, 55, 57, 80]. Heterologous vaccination (AZ ChAdOx1 followed by an mRNA vaccine) show highest protection [57]. Cases of SARS-CoV-2 infection despite complete vaccination (hereonafter referred to as breakthrough infection, BTI) have become very frequent since the emergence of the Omicron variants [33, 45-47, 56, 57]. Accordingly, *in vitro* neutralization assays highlight poor cross-neutralization of Omicron by sera from vaccinees [19, 32-36, 48, 68, 81, 82]. Although booster vaccine dose(s) restore neutralizing antibody (NAb) responses against Delta and partially against Omicron, they do not decrease viral load or transmission [35, 36, 44, 46, 48, 57, 81-90].

Protection against Omicron conferred by infection with other VOCs is still controversial. Some authors report that reinfection rates with Omicron are far superior to reinfection rates with Beta and Delta [91] and that NAbs elicited by pre-Omicron VOCs or by variant-specific vaccines are lower or ineffective against Omicron [27, 48, 68, 88, 92]. Others in contrast report that breakthrough infection with VOCs elicits cross-reactive antibodies with the ability to neutralize Omicron [49, 93, 94]. Converging reports indicate that Omicron BA.1 breakthrough infection elicits antibodies with neutralizing activity against Omicron BA.1 and previous VOCs [27, 48, 95-97] but poorly cross-neutralizing other Omicron sublineages [98]. In unvaccinated individuals, Omicron infections trigger poorly neutralizing antibodies against all VOCs [49, 95, 96]. The contribution of vaccine- or infection-induced immunity to the lower pathogenicity of Omicron and the long-term impact of infection by Omicron are difficult to set apart [14, 24, 26, 27, 95]. Nevertheless, the inverse correlation between antibody levels at the time of infection and viral load [27, 99] together with the high rate of hospital admissions recorded in Hong-Kong, where vaccine coverage is low, suggest that pre-existing immunity plays a major role in protection. The widespread dissemination of Omicron calls for a better understanding of its sensitivity to neutralization by antibodies induced by vaccination and prior infection.

Because of its spectacular infectivity, the Omicron wave caused a substancial number of breakthrough infections and reinfections, and in some cases, even co-infections. These are fertile ground for the surge of recombinants, which further complexify the epidemic landscape. Understanding the role of pre-existing immunity in this setting is thus fundamental. In this study we compared the ability of convalescent sera from 70 unvaccinated patients infected with the ancestral B.1 strain and from 16 BTI cases infected with Gamma (2 patients) or Delta (14 patients) to neutralize Omicron BA.1, using replicating viral strains isolated from patients in Luxembourg. Pre-VOC convalescent sera showed substantial heterogeneity in their cross-neutralizing ability against pre-Omicron VOCs and most failed to neutralize Omicron. Patients with moderate disease had overall higher neutralizing levels against all strains than patients with mild/asymptomatic disease or with severe or critical disease. In contrast, half the sera from BTI patients had high neutralizing ability against all tested strains, while half had no or low NAbs against all strain. Neutralizing BTI sera retained some ability to neutralize Omicron. Importantly, BTI sera had lower antibody levels than unvaccinated convalescent sera, suggesting higher affinity conferred by vaccination and hybrid immunity.

## Materials and Methods Patient samples

Serum from 70 unvaccinated patients (hereafter ‘convalescent sera’) who were infected between March and July 2020 and serum from 16 vaccinated patients with breakthrough infection (2 Gamma and 14 Delta) (hereafter ‘BTI sera’) who were infected between July 15th and September 20th 2021 were analyzed. All patients had RT-PCR-confirmed SARS-CoV-2 infection. The study was approved by the LIH Institutional Review Board (study number 14718697-NeutraCoV). Anonymized patient left-over samples collected at the Centre Hospitalier de Luxembourg (CHL) were used for the set-up of serological and virological tests in agreement with GDPR guidelines. No clinical data was available other than the time since symptom onset and the degree of disease severity recorded by the clinician for the unvaccinated patients. Disease severity stratification was as follows : *Mild/asymptomatic* patients (8 patients) presented flu-like symptoms or no symptoms; patients with *Moderate* disease (19 patients) had fever, flu-like symptoms, anosmia, fatigue, gastro-intestinal disturbances, but did not require hospitalization or oxygen supplementation; patients with *severe* or *critical* disease (43 patients) were admitted to the hospital, required oxygen supplementation and/or intensive care. For BTI cases, data on the lineage of the infecting strain and the time since 2^nd^ vaccine dose were provided by CHL. Convalescent sera were collected during acute infection (Median 14 days, IQR 9-20), while the BTI sera were collected at the time of diagnosis but time since symptom onset is not known.

### Cells

Vero-E6 cells (a kind gift from Dr. Thorsten Wolff, Influenza und respiratorische Viren, Robert Koch Institute, Germany) were maintained in DMEM supplemented with 10% Foetal Bovine Serum (FBS), 2 mM L-Glutamine, 50 µg/mL Penicillin and 50 µg/mL Streptomycin (all from Invitrogen, Merelbeke, Belgium). For infection experiments, 2% FBS was used (hereafter referred to as Viral Growth Medium, VGM).

### Serology

The MesoScale Diagnostics (MSD) V-Plex COVID-19 Coronavirus Panel 1 serology kit (K15362U) was used according to the manufacturer’s recommendations to determine the IgG profile of sera (MesoScale Diagnostics, Rockville, MD, USA). This multiplex assay includes SARS-CoV-2 antigens (N, S, RBD, NTD) as well as Spike proteins from other Coronaviruses (SARS-CoV, MERS-CoV, OC43, HKU1) and Influenza A Hemagglutinin H3.

### Virus isolation and titration

SARS-CoV-2 strains D614G and VOCs (Gamma, Delta and Omicron) were isolated from anonymized left-over patient nasopharyngeal swabs (NPS) collected from patients at the CHL. For isolation, 500 µl of residual swab preservation medium was added to Vero-E6 cells (1.2×10^6^ cells) and cytopathic effect (CPE) was monitored visually. Viral supernatant was used to constitute a viral stock by infecting Vero-E6 cells in a second passage. The viral supernatant from passage 2 was centrifuged and stored at -80°C until use. All experiments were performed with the same viral stock. Viral strains present in the orignial material (swabs) were identified through next-generation sequencing and the Spike was resequenced after the second passage to verify sequence conservation. We isolated representative strains for B.1 (D614G, pre-VOC), Gamma, Delta and Omicron (sublineage BA.1).

The 50% Tissue Culture Infectious Dose (TCID50) was assessed by titrating viral strains on Vero-E6 cells in sextuplicate wells. Briefly, 10^4^ cells/ well in a 96-well microtitre plate were infected with serial 10-fold dilutions of isolated virus for 72 hours at 37°C with 5% CO^2^. Virus-induced CPE was measured using the tetrazolium salt WST-8, which is cleaved to a soluble strongly pigmented formazan product by metabolically active cells (CCK-8 kit, Tebu-Bio, Antwerp, Belgium).

### Live-virus neutralization Assay

Serial two-fold dilutions of heat-inactivated patient serum were incubated 1 hour with 100 TCID50 of virus in VGM before inoculation of Vero-E6 cells (10^4^ cells/ well in a 96-well microtitre plate). Cells were cultured for another 72 hours at 37°C with 5% CO_2_. A positive control (no serum) and an uninfected control (no serum no virus) were included in each plate to assess maximum and minimum values. Virus-induced CPE was measured using the tetrazolium salt WST-8 as above. All infections were performed in triplicate wells. Percent survival was calculated relative to uninfected cells and the half-maximal inhibitory concentration for serum (IC50) was determined by inferring the 4-parameter non linear regression curve fit (GraphPad Prism v5). The top and bottom values were unconstrained.

### Statistical analyses

Statistical analyses were performed using GraphPad Prism v5. The Shapiro-Wilk test was used to verify distribution. Differences between groups were compared using a Mann-Whitney U test for comparison between two groups and a Kruskal-Wallis signed-rank test followed by a Dunn’s post-hoc test for multiple comparisons since most datasets were non-normally distributed. Correlation coefficients (r) were determined using as Spearman’s rank correlation. P-values < 0.05 were considered significant.

## Results

### Cross-Neutralization of Gamma, Delta and Omicron by pre-VOC convalescent sera

First, we measured the neutralizing ability of sera from 70 patients infected early in the pandemic with the ancestral pre-VOC D614G (B.1) strain. The sera were collected during acute infection (median 14 days, IQR 9-20) and most were from patients with moderate or severe/critical COVID-19. The half-maximal inhibitory concentrations (IC50) of convalescent sera against B.1, Gamma and Delta spanned a broad neutralization range and had similar geometric means (p> 0.05): IC50 B.1 = 0.007783, IC50 Gamma = 0.007653 and IC50 Delta = 0.007137 (Fig 1A and 1B and Supplementary Fig 1A-1C). One third (24/70, 34.3 %) of the convalescent sera failed to neutralize B.1, and a slightly but significantly higher proportion failed to neutralize Gamma (25/61, 41%, p<0.01) and Delta (29/67, 43.3%, p<0.01) at a 1:40 serum dilution. In contrast, all but one serum failed to neutralize Omicron at the highest serum concentration tested (57/58, 98.2%, p< 0.0001) and the geometric mean IC50 was significantly higher (IC50=0.04676, p<0.0001) (Fig 1C and 1D). Aside from Omicron, neutralizing capacities of convalescent sera varied depending on the variants. Sera with low or no neutralizing activity against one strain often showed higher neutralizing activity against another strain (Fig 1A-1C and 1E and Supplementary Fig 1A-1C). Accordingly, there was a good but not perfect correlation between neutralizing activities of convalescent sera against B.1 and Gamma (Spearman’s r = 0.6852, p<0.0001) or Delta (Spearman’s r = 0.6924, p<0.0001) (Fig 1E). The highest neutralizing activity against B.1 and VOCs was achieved by sera from patients with moderate disease (Fig 1F), thus highlighting the plasticity of the antibody response vis-à-vis different variants. The time elapsed between symptom onset and serum collection contributed only marginally and not significantly to the differences in neutralizing ability against B.1 (Supplementary Fig 1D).

**Figure 1.**
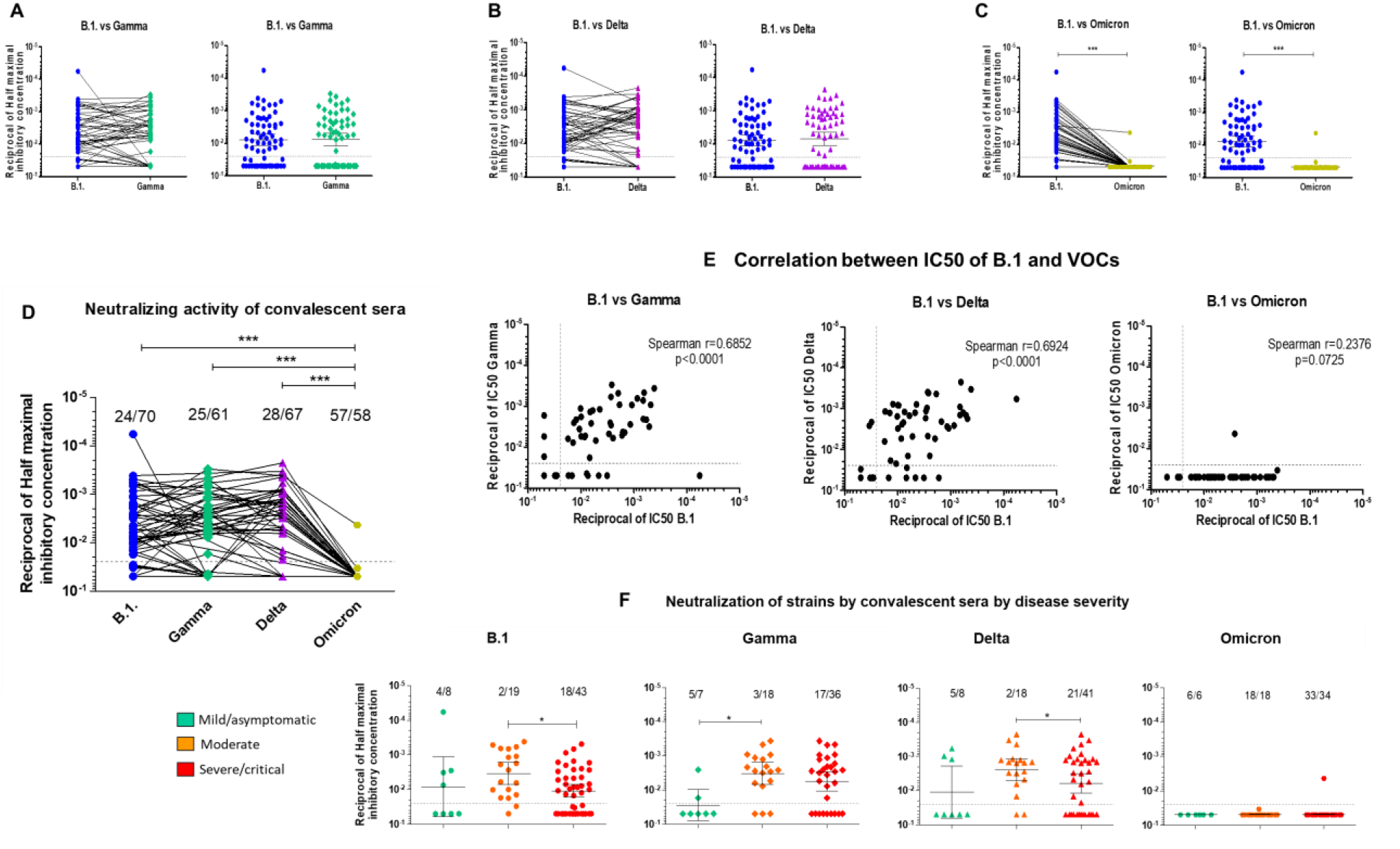
Neutralizing activity of pre-VOC unvaccinated SARS-CoV-2-infected convalescent sera against ancestral and VOC strains. **A-C. Pairwise comparison of half-maximal inhibitory concentrations (IC50) of B.1 with Gamma (A), Delta (B) and Omicron (C).** Serial two-fold dilutions (starting 1:40, grey dotted line on graph) of heat-inactivated convalescent sera were incubated 1 hour with 100 TCID50 of virus in Viral Growth Medium. Vero-E6 cells (10^4^ cells/ well) were infected with the mixture for 72 hours at 37°C. Virus-induced cytophathic effect (CPE) was measured using the tetrazolium salt WST-8. All infections were performed in triplicate wells. Percent survival was calculated relative to uninfected cells and the half-maximal inhibitory concentration for serum (IC50) was determined by inferring the 4-parameter non linear regression curve fit (GraphPad Prism v5). The top (100% survival) and bottom (no serum) values were unconstrained. **D. Comparison of IC50 for all strains**. The proportion of non-neutralizing sera is indicated above each data set. **E. Correlation between the IC50 of B.1 and VOCs. F. IC50 of convalescent sera against B.1 and VOCs for patients with mild/asymptomatic (green), moderate (orange) or severe/critical (red) forms of COVID-19**. For all analyses, differences between groups were compared using a Mann-Witney U-test for comparison between two groups and a Kruskal-Wallis followed by a Dunn’s multiple comparison post-hoc test when three or more groups were compared. P-values < 0.05 were considered significant. *: p<0.05; **: p<0.01; ***: p<0.001.

To gain some insight into the target and efficacy of antibodies mediating the neutralizing activity, we measured antibodies targeting the viral Spike (S), the Receptor Binding Domain (RBD) and the S N-terminal Domain (NTD) as well as antibodies targeting the viral Nucleocapsid (N) using the MSD V-plex platform. All patients had detectable antibodies against N, S and the RBD, ranging over 5 orders of magnitude (Fig 2A). Anti-NTD antibody levels were lower, and undetectable in a handful of patients with severe disease (Fig 2A). Antibody levels only moderately correlated with neutralizing ability (Spearman r=-0.5522 for N, Spearman r=-0.6537 for S, Spearman r =-0.6301 for RBD and Spearman r=-0.4923 for the NTD, p<0.0001) (Fig 2B). In line with neutralizing ability, patients with moderate disease had significantly higher antibody levels against all viral determinants (N, S, RBD ant NTD) than patients with severe disease forms (Fig 2C).

**Figure 2.**
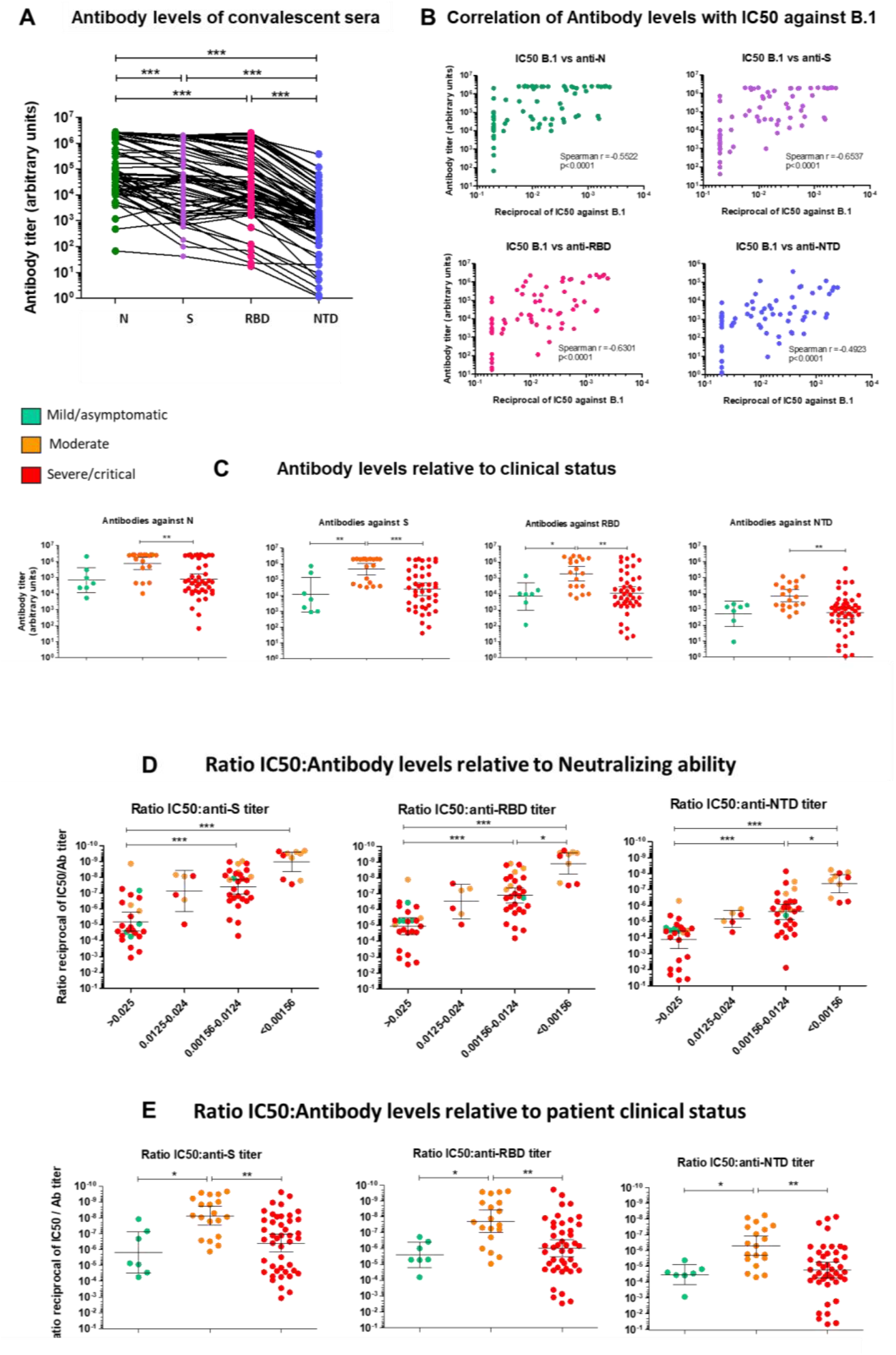
Serological analysis of convalescent sera. **A. Antibody levels in convalescent sera.** Antibody levels against SARS-CoV-2 N (green), S (purple), the Receptor Binding Domain (RBD) (pink) or the N-terminal domain (NTD) (blue) of Spike were measured in convalescent sera using the MSD V-plex platform for SARS-CoV-2. Antibody levels are reported as arbitrary units. **B. Correlation between half-maximal inhibitory concentration for serum (IC50) and antibody levels against SARS-CoV-2 N, S, RBD and NTD. C. Antibody levels of convalescent sera startified by disease severity. C and D. Ratios of IC50 to antibody levels against S, RBD and NTD stratified according to neutralizing activity (D) or to patient clinical status (E)**. For all analyses, differences between groups were compared using a Mann-Witney U-test for comparison between two groups and a Kruskal-Wallis followed by a Dunn’s multiple comparison post-hoc test when three or more groups were compared. P-values < 0.05 were considered significant. *: p<0.05; **: p<0.01; ***: p<0.001.

We further assessed neutralization activity in regard of anti-S, anti-RBD and anti-NTD antibody levels (ratio IC50: Antibody levels). This ratio can be used as a proxy for antibody affinity [100]. As shown in Fig 2D, the IC50:Ab level ratio decreased markedly with increasing neutralizing ability for all antibodies (p<0.001 for sera with IC50s < 0.0124 compared to non-neutralizing sera). Overall, the IC50:S and IC50:RBD ratios were similar, while the IC50:NTD ratio was ∼1 to 1.5 log_10_ higher, reflecting the fact that antibodies targeting the RBD represent the most abundant fraction of antibodies targeting S and account for most of the neutralizing ability. Again, the lowest ratio (highest neutralizing activity and highest Ab levels) was achieved by sera from patients with moderate disease (Fig 2E), suggesting not only a quantitative increase in NAb, but also an increase in their affinity against S.

### Cross-Neutralization of sera from vaccinated breakthrough infection (BTI) cases

Next, we assessed the ability of sera from 16 vaccinated individuals with breakthrough infection (BTI) with Gamma (2 cases) or Delta (14 cases) to neutralize the ancestral B.1 strain, the corresponding infecting VOC (Gamma or Delta) and Omicron BA.1. BTI were infected between July 15^th^ and September 18^th^ 2021, when Gamma and Delta were the main circulating VOCs in Luxembourg. The median time elapsed since the 2^nd^ vaccine dose was 3.06 months [CI=2.09-4.47]. Most sera were collected at the time of diagnosis but time since symptom onset is not known. Typically, BTI sera could be categorized into two clearly distinct categories, non-neutralizing or very poorly neutralizing sera (only at highest serum concentration tested, 1:40) (8 sera) or highly neutralizing sera (8 sera) (Fig 3A). Among the non-neutralizing sera, 5 (4 Delta, 1 Gamma) had no neutralizing ability against B.1 at the highest serum concentration tested (1:40). Geometric mean IC50 was slightly but significantly lower against Delta (Geometric mean IC50=0.002887) than against B.1 (Geometric mean IC50=0.004839) (p<0.05) (Fig 3A), indicating better neutralizing ability against the infecting strain than against the wild-type virus the vaccines are designed on. Geometric mean IC50 could not be calculated for the two BTI infected with Gamma, but they also fell into one of the two profiles, one having no neutralizing ability and one with good neutralizing ability (Fig 3A). In contrast to what was recorded for convalescent sera, the neutralizing profile was maintained across strains overall, such that all sera which neutralized B.1 at dilutions higher than 1:80 retained their neutralizing ability against the corresponding VOC or had higher neutralizing ability against VOCs. Vice versa, most sera which did not neutralize B.1 also failed to neutralize the infecting VOC (1 Gamma and 3 of 4 non-neutralizing Delta BTI) (Fig 3A). Therefore, while it is easily conceivable that individuals with no or low neutralizing ability would be susceptible to breakthrough infection, more than half the BTI cases had NAbs against Delta (or Gamma). Furthermore, their sera cross-neutralized Delta better than the ancestral B.1 strain, arguing against the hypothesis that immune escape underlies breakthrough infection in these cases.

**Figure 3.**
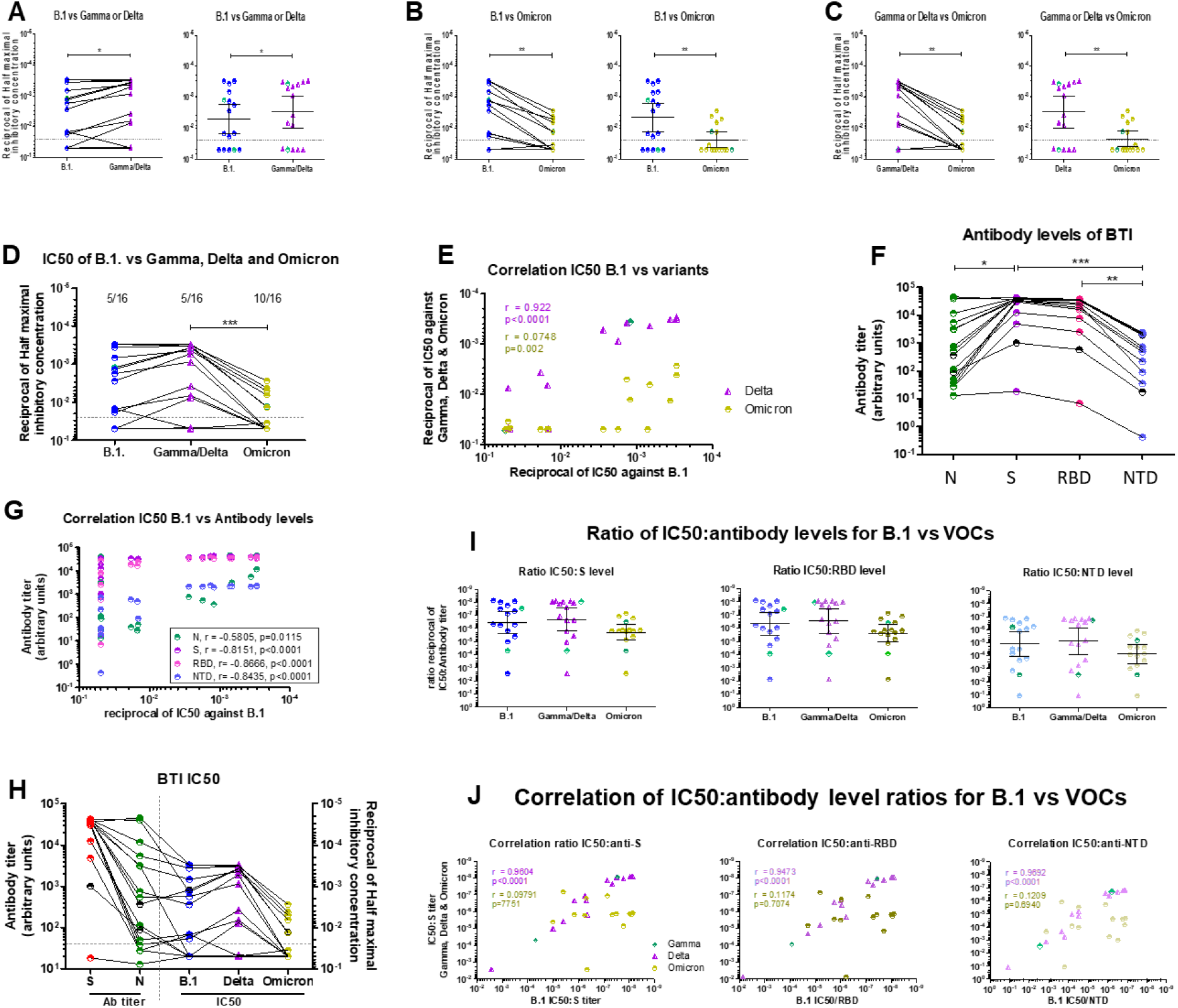
Neutralizing activity and serology of sera from vaccinated breakthrough infections (BTI) against B.1, Gamma, Delta and Omicron BA.1. **A-C. Pairwise comparison of Neutralization of B.1, Gamma, Delta and Omicron.** Serial two-fold dilutions (starting 1:40, grey dotted line) of heat-inactivated sera from breakthrough infection cases (BTI) collected around the time of infection were incubated 1 hour with 100 TCID50 of representative ancestral D614G (B.1), Gamma, Delta or Omicron BA.1 SARS-CoV-2 strains in Viral Growth Medium. Vero-E6 cells (10^4^ cells/ well) were infected with the mixture for 72 hours. Virus-induced CPE was measured using the tetrazolium salt WST-8. All infections were performed in triplicate wells. Percent survival was calculated relative to uninfected cells and the half-maximal inhibitory concentration for serum (IC50) was determined by inferring the 4-parameter non linear regression curve fit (GraphPad Prism v5). The top (100% survival) and bottom (no serum) values were unconstrained. IC50 or Gamma and Delta are grouped and are represented in green diamonds and purple triangles respectively **D. Comparison of IC50 for all strains**. The proportion of non-neutralizing sera is indicated above each data set. **E. Correlations between half-maximal inhibitory concentration (IC50) of B.1 and Gamma (green diamonds), Delta (purple triangles) and Omicron BA.1 (gold circles). F. Serology of BTI sera**. Antibodies against SARS-CoV-2 N (Nucleocapsid), S (Spike), the Spike RBD (Receptor-Binding Domain) or the Spike NTD (N-terminal Domain) were measured using the MSD V-Plex COVID-19 Coronavirus Panel 1 serology kit. Antibody levels are reported as arbitrary units. Sera from BTI infected with Gamma are in black, those infected with Delta are colored. **G. Correlation between IC50 and antibody levels. H. Anti-S and anti-N antibody levels (Left Y-axis) and half maximal inhibitory concentrations of BTI sera against B.1, Gamma/Delta and Omicron (Right Y-axis)**. Gamma BTI are highlighted in black in all cases. **I. Ratios of IC50 to B. (blue circles), Gamma (green diamonds), Delta (purple triangles) and Omicron (gold circles) divided by antibody levels against S, RBD and NTD in BTI sera. J. Correlations between ratios of IC50 to antibody levels against S, RBD and NTD in BTI sera**. Color code: B.1: blue circles; Gamma: green diamonds, Delta: purple triangles, Omicron BA.1: gold circles. For all analyses, differences between groups were compared using a Mann-Witney U-test for comparison between two groups and a Kruskal-Wallis followed by a Dunn’s multiple comparison post-hoc test when three or more groups were compared. P-values < 0.05 were considered significant. *: p<0.05; **: p<0.01; ***: p<0.001.

Next, we assessed the cross-neutralizing ability of these sera against Omicron BA.1. As shown in Fig 3B-3D, all BTI sera featured a substantial drop in neutralizing ability against Omicron (Geometric mean IC50 = 0.02358, p<0.01 compared to B.1 and to Delta). However, of the 8 BTI sera with high neutralizing ability against B.1 and Gamma or Delta, 6 also retained partial neutralizing ability against Omicron (Fig 3B-3D). Accordingly, there was a very good correlation between the IC50s of BTI sera against B.1 and Gamma or Delta (Spearman r = 0.922, p< 0001), but not between B.1 and Omicron (Spearman r=0.0748, p=0.002) (Fig 3E).

All but one BTI sera had high antibody levels against S, including the RBD and the NTD (Fig 3F), in line with them being vaccinated. In contrast, antibodies targeting N spanned a broad range (Fig 3F), suggesting infection may have been going on for some days before diagnosis in some BTI patients. Neutralizing ability against B.1 correlated nicely with antibodies against S, the RBD and the NTD (Spearman r =-0.8151, p<0.0001 for S, r=-0.8666, p<0.0001 for RBD and r=-0.8435, p<0.0001 for the NTD) and less with antibodies against N (Spearman r=0.5805, p=0.01115) (Fig 3G), confirming that breakthrough infection does not reflect a lack of circulating NAbs directed against Spike. Higher anti-N antibodies were present in sera with higher neutralizing ability (Fig 3H). This observation coupled to the fact that patients with high antibody levels against S targets did not necessarily have high anti-N antibody levels suggests either different time elapsed since infection or a lag in the onset of the antibody response in some BTI cases.

The IC50:Ab level ratio was highest for S and the RBD and ∼1.5 log_10_ lower for antibodies targeting the NTD, suggesting that most anti-S antibodies elicited by vaccines target the RBD, while antibodies targeting the NTD are much less abundant (Fig 3I). The IC50:Ab level ratio was similar for B.1 and Gamma/Delta for all antibdoes targeting S (Fig 3J), reflecting the high neutralizing ability of antibodies against the infecting virus. The IC50:Ab level ratio against Omicron was nearly 1 log_10_ higher (Fig 3F), again reflecting the lower efficacy of antibodies against this variant.

These figures highlight differences in the cross-neutralizing ability of convalescent sera in comparison to BTI sera. The former showed different cross-neutralization profiles against Gamma and Delta VOCs and retained no neutralizing ability against Omicron, while BTI sera showed more similarities across variants and only a partial loss in neutralization against Omicron.

### Comparison of BTI and convalescent sera

When compared to convalescent sera, BTI sera had higher neutralizing activity against B.1 as well as against VOCs, including Omicron(Fig 4A). This efficient neutralizing ability allowed to cross-neutralize Omicron, as testified by the observation that 6/8 BTI sera with neutralizing activity against B.1 and Gamma/Delta partly neutralized Omicron. This result clearly indicates that breakthrough infections with pre-Omicron VOCs induce an antibody response which cross-neutralized Omicron BA.1, in contrast to convalescent sera. Remarkably, the higher neutralizing activity was achieved with significantly lower antibody levels against N, S, the RBD and the NTD (p<0.0001 in all cases) in BTI sera compared to convalescent sera (Fig 4B), indicating that higher neutralizing efficacy is reached with lower antibody titers, thus hinting at a greater affinity of antibodies. The neutralizing activity remained significantly higher (p<0.001) when only patients with moderate infection, which display the highest neutralizing activity, were compared to BTI (not shown). Accordingly, the IC50:Ab level ratios were comparable for BTI and convalescent sera (Fig 4C), confirming that the lower IC50 (higher neutralizing ability) necessitated lower antibody titers. This observation held true for all VOCs and for anti-S, anti-RBD and anti-NTD antibodies. Convalescent sera from patients with moderate disease have higher neutralizing ability (Fig 1F) and higher antibody levels (Fig 2E) than convalescent sera from patients with mild/asymptomatic disease or with severe or critical disease. Nevertheless, the IC50:Ab levels ratio of patients with moderate disease was lower than that of BTI (Fig 4D), indicating that higher neutralizing ability (lower IC50) necessitated higher antibody levels. Therefore, antibodies elicited by vaccines combined to infection, although much less abundant than those elicited by infection during the acute phase, seem to have higher affinity than those elicited by infection alone, even in individuals with efficient neutralizing responses and contained disease manifestations. This may reflect a more targeted humoral response induced by vaccination over natural infection, or the superiority of hybrid immunity over immunity elicited by infection alone [93, 100, 101]. The observation that BTI patients with higher anti-N antibody levels had the highest neutralizing activity argues in favor of the second hypothesis.

**Figure 4.**
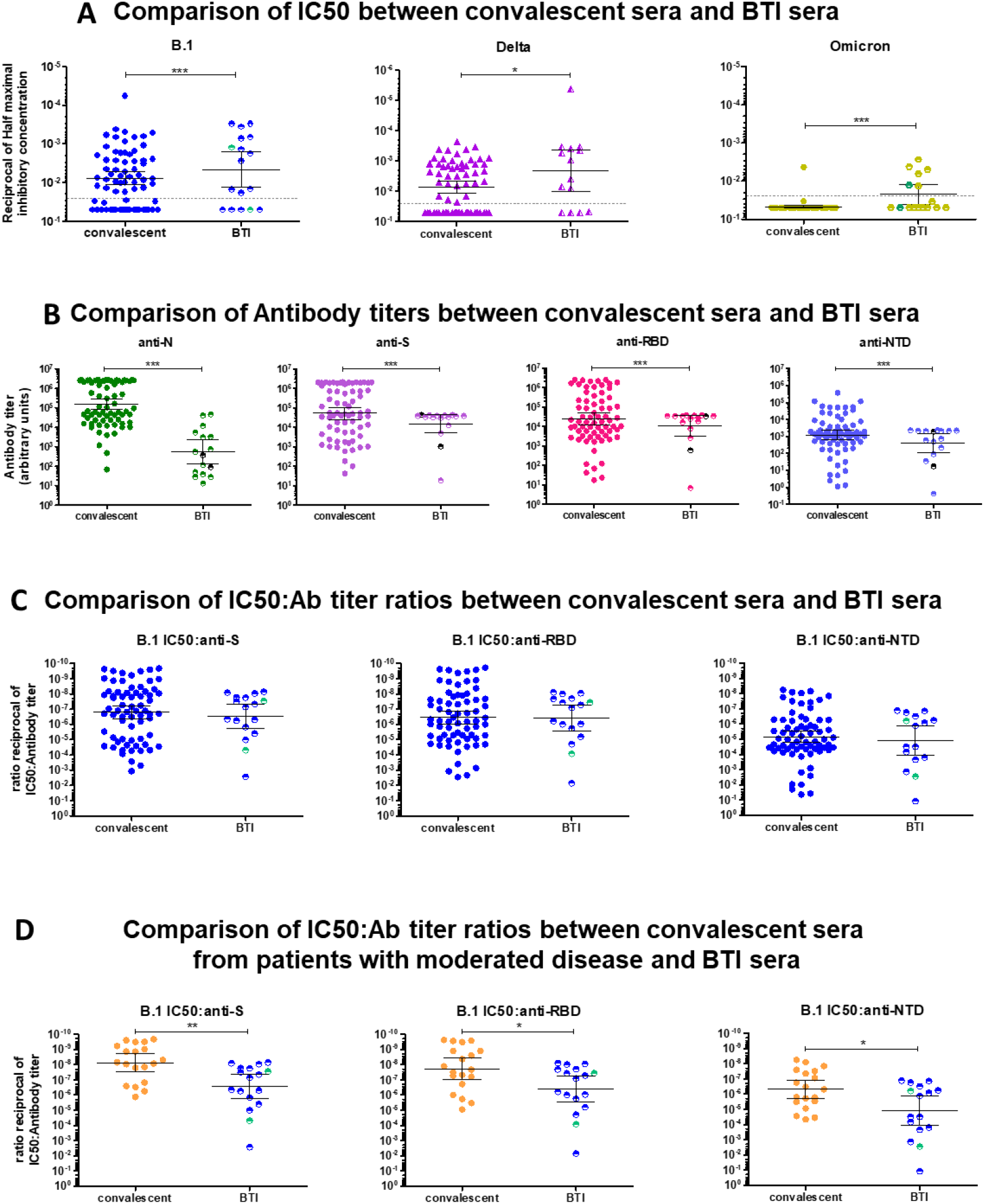
Comparison of neutralizing activities of BTI and convalescent sera against B.1, Gamma, Delta and Omicron. **A. Comparison of half-maximal inhibitory concentrations (IC50) of convalescent and BTI sera against B.1, Delta and Omicron.** For BTI sera, the patients infected with Gamma are represented with green circles. A pairwise comparison with Gamma was not possible since only two BTI infected with Gamma were available. The grey dotted line represents the 1:40 serum dilution cut-off. **B. Comparison of antibodies against N, Spike (S), RBD and NTD in BTI and convalescent sera**. For BTI sera, the patients infected with Gamma are represented with black circles. **C. Comparison of the B.1 IC50:antibody level ratios for BTI and convalescent sera**. For BTI sera, the patients infected with Gamma are represented with green circles. **D. Comparison of the B.1 IC50:antibody level ratios for BTI and convalescent sera with moderate disease**. For BTI sera, the patients infected with Gamma are represented with green circles. For all analyses, differences between groups were compared using a Mann-Witney U-test for comparison between two groups and a Kruskal-Wallis followed by a Dunn’s multiple comparison post-hoc test when three or more groups were compared. P-values < 0.05 were considered significant. *: p<0.05; **: p<0.01; ***: p<0.001.

## Discussion

In this study we compared the neutralizing abilities and antibody levels of sera from unvaccinated individuals infected with the ancestral B.1 (D614G) strain (convalescent sera) and from vaccinated BTI individual. Overall, ∼30% of convalescent sera and 50% of BTI sera were non-neutralizing against B.1 and VOCs Gamma and Delta (Fig 1A-1D and Fig 3A-3D). Both convalescent and BTI sera retained good neutralizing ability against Gamma and Delta and displayed a substantial decrease in neutralizing ability against Omicron (Fig 1A-1D and Fig 3A-3D) in agreement with previous studies [32-36, 45-48, 56, 57, 68, 72-79, 81, 96, 100, 102-105]. It is thought that the ability of Omicron to elude NAbs reflects a combination of higher affinity of Spike for ACE2 [21, 29, 30], together with TMPRSS-2-independent, endocytosis-mediated entry [21, 106, 107]. Furthermore, the Omicron lineage is phylogenetically and serologically distinct from the other SARS-CoV-2 lineages. The Omicron Spike also adopts a distinct more compact conformation and glycosylation patterns that allow it to evade NAbs [31]. The fact that prior infection and vaccination do not fully protect against infection with Omicron or symptomatic disease but do partly protect against severe disease [108] suggests that T-cells and memory B-cells, as well as other innate immune signatures [109] are starring.

Although both convalescent and BTI sera were immunized against the ancestral pre-VOC strain, either through natural infection or through vaccination, we recorded qualitative differences within and between both groups of patients. First, the cross-neutralizing ability of convalescent sera was markedly strain/variant-dependent, with some sera being non-neutralizing against B.1 but neutralizing against Gamma or Delta (Fig 1A-1D and Supplementary Fig 1A-1C). Furthermore, there was substantial heterogeneity in the neutralizing ability and antibody levels among unvaccinated convalescent patients. Patients with moderate disease generally had the highest antibody levels (Fig 2A and 2C) and the highest neutralizing activity against B.1 as well as against pre-Omicron VOCs (Fig 1F), while the majority of convalescent sera which failed to neutralize B.1 were from patients with mild disease, where innate immunity sufficed to clear infection, or from patients with severe or critical disease (Fig 1F). The correlation and predictive power of IgG antibody levels with disease severity is controversial, as some studies report that antibody levels and neutralizing activity parallel disease severity [110-113] while others find higher anti-Spike antibody levels in non-ICU patients [114]. Our results (Fig 2C and 2D) agree with the latter study, and reinforce the importance of the potency and affinity of neutralizing antibodies in viral clearance and protection against severe disease. Accordingly, this distinction between groups of patients was sharper when the IC50:Ab level ratio was calculated (Fig 2E). This ratio was suggested to provide insight into the affinity of antibodies for Spike [100, 113, 115] and thus highlights that the antibody response mounted by patients with severe disease is generally poorly neutralizing and may be ‘unfocused’. While the IC50:Ab level ratio of patients with moderate disease was lower than that of patients with servere or critical disease, it remained lower than that of BTI patients (Fig 4D), suggesting that unvaccinated patients required higher antibody levels to achieve similar neutralization. Sera from BTI patients, in contrast, had a much more uniform cross-neutralizing pattern, as they were either non-neutralizing or neutralizing against the ancestral B.1 strain and the infecting VOC (Fig 3A-3D). Neutralizing activity of BTI sera correlated very well with antibody levels (Fig 3G), while the correlation was more modest for convalescent sera (Fig 2B), highlighting qualitative differences in antibodies elicited by vaccines and by infection. Vaccination or hybrid immunity lead to higher neutralizing ability and broader cross-neutralization spectrum (Fig 4A), despite lower overall antibody levels (Fig 4B and 4C), reflecting higher antibody affinity. Notably, most BTI patients had high antibody levels against S and the RBD and high neutralizing activity against B.1, Gamma and Delta, i.e. against their infecting VOC, indicating that breakthrough infection occurred despite the presence of NAbs. One possible explanation is that the increased affinity and faster binding of variant Spikes to ACE2 [4, 21, 29, 30] may short-circuit NAbs in some patients. Alternatively, because we measured the presence of NAbs in serum, NAbs may be scarce at the site of infection (upper respiratory airways), allowing infection. These antibodies may however play a role in protecting against severe disease and death.

Second, neutralizing BTI sera partially retained neutralizing capacity against Omicron (Fig 3B-3D), while only one convalescent serum was able to neutralize Omicron (Fig 1C and 1D). In our study, the proportion of BTI patients with neutralizing antibodies against Omicron above the dilution cut-off of 1:40 was 37.5% (6/16, 5 Delta and 1 Gamma), which is less than that reported by a previous study (∼75%) [105]. The ability of Delta-elicited sera to cross-neutralize Omicron is debated [48, 49, 68, 88, 91-94, 116]. Our results clearly indicate that Gamma or Delta-breakthrough sera with high neutralizing activity against these VOCs are able to cross-neutralize Omicron BA.1, in agreement with previous studies [49, 93, 94, 117], although our data apply to vaccinated BTI infected with Delta and we cannot infer whether they extend to BTIs infected with other VOCs. Since breakthrough infection acts as a booster, this observation is congruent with the restoration of neutralizing ability induced by booster vaccination [35, 36,44,46,48,57,81-89]. It is currently acknowledged that vaccination of recovered COVID-19 patients with mRNA vaccines (hybrid immunity) induces IgG titers similar to or higher than two doses of vaccines in SARS-CoV-2-naïve individuals [27, 118, 119] and protects better against Omicron BA.1 than vaccination alone [49]. Conversely, Omicron BA.1 infection alone offers limited protection against pre-Omicron and against Omicron BA.2-related strains, while breakthrough infection after vaccination confers some cross-protection [95]. These results are in line with the observation that Omicron BA.1 variant-based vaccines including Delta determinants have higher neutralizing ability against Omicron sublineages compared to vaccines which include only Omicron determinants [116, 120]. They also indirectly confirm the serologic similarities between B.1 and Delta (Fig 1B and 1D and Fig 3A, 3D and 3E) on one hand and the serotypic specificity of the Omicron lineage and sublineages on the other. Hybrid immunity appears to be more durable and seems to cross-protect more efficiently than infection of vaccination individually [27,101,119,121-123], likely as a result of antibody affinity maturation [67, 100, 124]. In our case, BTI were not followed in time and the time elapsed since infection is not known. Nevertheless, the observation that patients with high anti-N antibody levels (> 1000 arbitrary units) had higher neutralizing ability (Fig 3H) suggests a longer time window since infection. Affinity maturation proceeds in infected patients in secondary lymphoid organs long after viral clearance from the upper respiratory airways [115, 125-129], as well as after vaccination [67,94,100,105,124,130]. Further supporting the importance of ongoing affinity maturation, breakthrough infection with Delta or Omicron after full vaccination with mRNA vaccines (2 doses), and more specifically the time window between the second vaccine dose and breakthrough was reported to elicit cross-reactive antibodies effective against Omicron [49, 93, 100]. Altough we could not confirm the role of time between vaccination and breakthrough in this study, it is possible that the time elapsed since vaccination (∼3 months) and the combination of different strains (Wuhan-based vaccines + infection with Gamma or Delta) may play a role. One recent report comparing neutralization of Omicron sublineages BA.1, BA.2, BA.4 and BA.5 by sera from infected-vaccinated and BTI individuals further shows that sera from BTI are much more neutralizing than sera from infected-vaccinated individuals [19]. This observation probably reflects the combination of new antibody responses neutralizing similar yet different strains together with affinity maturation of existing NAbs in BTI sera, while in infected-vaccinated sera, affinity maturation is “constrained” towards the ancestral strain. It is however noteworthy that the BTI patients in this study were infected with Omicron, while the BTI cases of our study were infected with pre-Omicron, phylogenetically more distant strains.

One shortcoming of this study is that it only includes a small number of BTI cases, particularly infected with Gamma. This reflects the relatively small number of breakthrough infections that required hospital admission. Despite these small figures, our results highlight discrete patterns among BTI and significant differences between unvaccinated-infected and BTI humoral responses. Another caveat is that we could not compare immune responses from unvaccinated-individuals and breakthrough infections infected with the same variant. Again, this situation ensues from the epidemic dynamic landscape, since vaccines were not available at the time the B.1 strain was circulating while VOCs had supplanted the ancestral strain after the roll-out of vaccine campaigns. Nevertheless, since most vaccines are designed based on the ancestral Wuhan stain, the epitopes targeted by antibodies elicited by infection and by vaccination should be comparable. Furthermore, the epidemic landscape on which the Omicron wave emerged is made of a multiplicity of past infections, different vaccines, vaccination times and patterns. As current vaccines are designed on the ancestral Wuhan strain and most BTI cases are due to Delta or Omicron, the data we report here is still representative of the grounds on which Omicron emerged. Moreover, while most individuals in Europe and the US have been vaccinated, in developing countries a large proportion of the population has not been vaccinated. In this context, prior immunity is only due to prior infection. Until now, VOCs have emerged in such a context, i.e. in a context of suboptimal neutralization. As increasing numbers of individuals become infected/reinfected or receive vaccines and boosters, SARS-CoV-2 variants will have to continue their evolutionary adaptation to overcome pre-existing immunity elicited by vaccines and by infections with distinct variants. It is thus possible that future variants will emerge from a completely distinct lineage, generating one or more serotypes [131]. Only those variants with transmissibility rates higher than Omicron and the ability to evade prior immunity will be able to supplant the current Omicron subtypes and recombinants. A close and rapid monitoring will be essential to assess their pathogenic potential and their susceptibility to NAbs elicited by previous VOCs. As Omicron variants are the main culprits for current BTI cases and elicit broad NAb responses [27], their cross-neutralizing ability will likely shape viral evolution. It will thus be essential to monitor the cross-neutralizing ability of Omicron-infected sera against emerging VOCs.

In conclusion, our data confirm and extend previous reports on the full immune escape of Omicron against neutralizing antibodies elicited by prior infection underline the protective role of hybrid immunity and the better cross-neutralizing ability conferred by infection in vaccinated individuals compared to unvaccinated individuals. Moreover, the overlap of Omicron sublineage-breakthrough infections has focused much research on the cross-neutralization within Omicron sublineages. Our data on the cross-neutralization of antibodies induced by of infection with pre-Omicron variants on Omicron BA.1 sheds light on the epidemic landscape on which Omicron spread. As new SARS-CoV-2 lineages with different serotypes may still emerge, a better understanding of the dynamics of humoral resposes will help appreciate the impact on the epidemic.

## Data Availability

All data produced in the present work are contained in the manuscript

## Author Contributions

DPB designed the study and acquired funding; MK, GG, TS and VA attended patients, managed and selected samples; ESS, JYS and DPB performed neutralization experiments; ESS, JYS, EC, CS and DPB isolated and characterized viral strains; OD, CH and MO contributed serological analyses. ESS and DPB analysed the data; ESS, DPB and CD drafted the manuscript.

### Funding

This work was supported by the Fonds National de la Recherche du Luxembourg (FNR COVID-19 FT-1 (**14718697** NEUTRACOV), by the Rotary Clubs Luxembourg, and by Ministère de l’Education et de la Recherche du Luxembourg.

### Institutional Review Board Statement

The study was approved by the LIH Institutional Review Board (14718697 NeutraCoV) and conducted in accordance with the Declaration of Helsinki.

### Informed Consent Statement

Patient consent was waived due to the the use of anonymized left-over samples for the validation of research tests in line with GPDR guidelines.

## Supplementary data

**Supplementary Figure 1.**
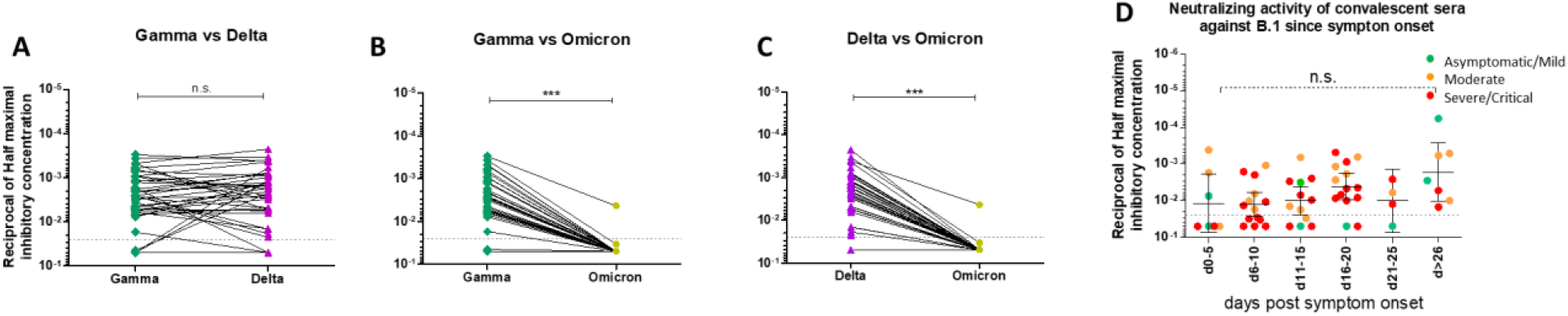
A-C. Pairwise comparison of half-maximal inhibitory concentrations (IC50) of convalescent sera against VOCs. Half-maximal inhibitory concentrations against each VOC were compared. Differences between groups were compared using Mann-Whitney test(A-C) or a Kruskal-Wallis followed by a Dunn’s multiple comparison post-hoc test (D). P-values < 0.05 were considered significant. *: p<0.05; **: p<0.01; ***: p<0.001. **D**. Neutralizing ability of sera against B.1. in relation to time since symptom onset for patients with mild/asymptomatic disease (green), moderate disease (orange) or severe/critical disease (red). The grey dotted line represents the 1:40 serum dilution cut-off.

**Supplementary Figure 2.**
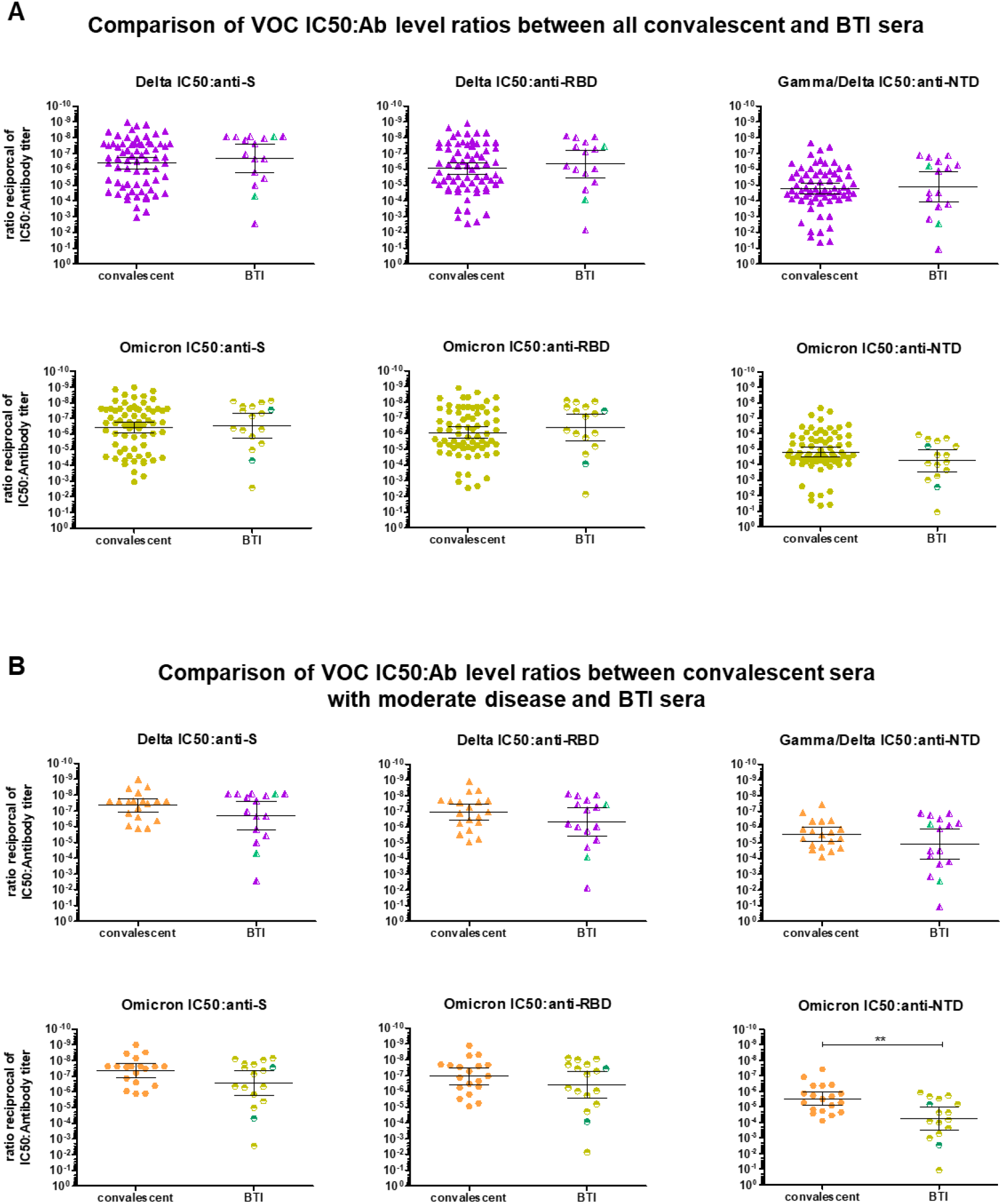
Comparison of the Delta (purple) and Omicron (yellow) IC50:antibody level ratios for BTI and convalescent sera for all convalescent patients (A) and for convalescent patients with moderate disease (B). For BTI sera, the patients infected with Gamma are represented with green circles. Differences between groups were compared using Mann-Whitney test(A-C) or a Kruskal-Wallis followed by a Dunn’s multiple comparison post-hoc test (D). P-values < 0.05 were considered significant. *: p<0.05; **: p<0.01; ***: p<0.001.

